# Improving prenatal detection of congenital heart disease with a scalable composite analysis of six fetal cardiac ultrasound biometrics

**DOI:** 10.1101/2024.08.13.24311793

**Authors:** Aneela Reddy, Sara Rizvi, Anita J. Moon-Grady, Rima Arnaout

## Abstract

Although screening of prenatal congenital heart disease (CHD) has improved over the last decade, the diagnosis rate can still be as low as 40%. The axial 4 chamber (A4C) is the most reliably obtained cardiac view in the fetal screening ultrasound but alone only has a maximum clinical sensitivity of 50-60%, particularly in large multicenter studies in low-risk populations. Standard biometrics, like cardiac axis (CA), cardiothoracic ratio (CTR) and cardiac chamber fractional area change (FAC), have individually been shown to be useful for CHD screening and can all be obtained from A4C alone. However, these biometrics are vastly underutilized because they are time-consuming to extract and difficult to interpret all at once. We hypothesized that using six standard biometrics in combination can improve complex CHD screening versus any one biometric alone. K-means clustering was performed to segregate the patterns of heart measurements into clusters. Sensitivity and specificity for CHD was 87% and 75%, respectively. Here, we demonstrate that a composite of six standard biometric has better sensitivity and accuracy for CHD than any one biometric alone and better than A4C visual assessment.

## MAIN TEXT

Although screening of prenatal congenital heart disease (CHD) has improved over the last decade, the diagnosis rate can still be as low as 40%^1^. The axial 4 chamber (A4C) is the most reliably obtained cardiac view in the fetal screening ultrasound but alone only has a maximum clinical sensitivity of 50-60%, particularly in large multicenter studies in low-risk populations.^2^ Standard biometrics, like cardiac axis (CA), cardiothoracic ratio (CTR) and cardiac chamber fractional area change (FAC), have individually been shown to be useful for CHD screening and can all be obtained from A4C alone.^3,4^ However, these biometrics are vastly underutilized because they are time-consuming to extract and difficult to interpret all at once.

We hypothesized that using six standard biometrics in combination can improve complex CHD screening versus any one biometric alone. We included 105 fetal echocardiograms (20 normal, 85 abnormal comprising 12 different CHD lesions). The chambers, thorax, and spine from the A4C view were segmented by an expert reader, and CA, CTR, right ventricular (RV) FAC, left ventricular (LV) FAC, right atrium:left atrium area ratio, and RV:LV area ratio were automatically calculated (Fig 1). K-means clustering (scikit-learn.org/) was performed to segregate the patterns of heart measurements into clusters.

**Figure 1.**
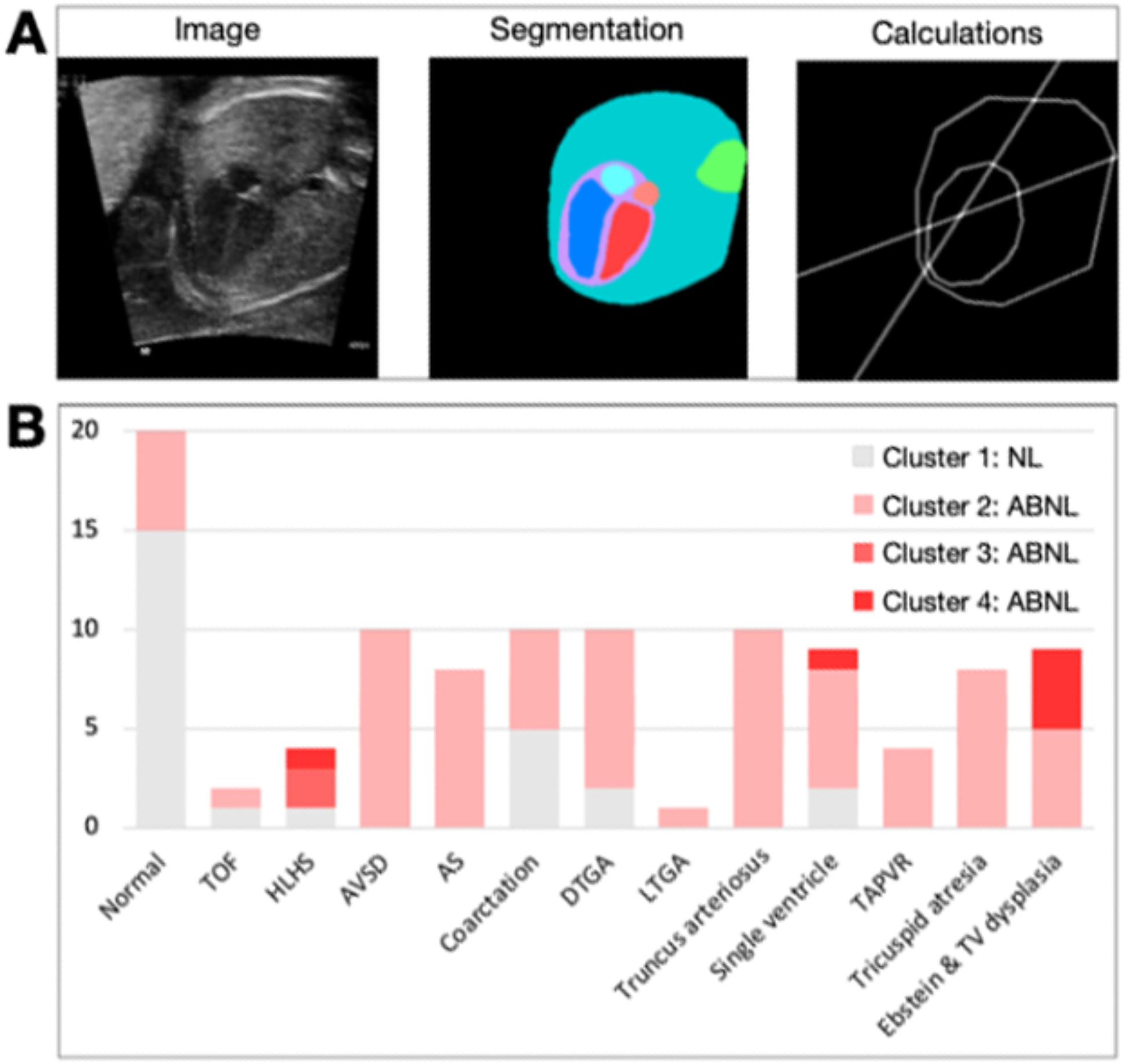
Segmentation and automatic calculation of biometrics. (A) Axial 4-chamber images were segmented (LV (red), RV (blue), LA (light red), RA (light blue), heart (purple), thorax (teal), and spine (green)). Chamber area ratios, cardiothoracic ratio, and cardiac axis measurements were automatically calculated from these segmentations; chamber fractional area changes were calculated using similar segmentations across the cardiac cycle. Calculations for fractional area change, RA:LA and RV:LV ratio were all derived from pixel areas. (B) K-means clustering of the resulting measurements is used as a composite biometric. Clustering result by lesion is shown.

The optimal number of clusters was four (based on silhouette score), with RV:LV ratio and CTR as the most important features distinguishing clusters. Cluster 1 was predominantly normal hearts. Cluster 2 consisted of lesions with either typically normal A4C views (aortic coarctation, d-transposition of great arteries (dTGA)) or more subtle abnormalities in the A4C view (tetralogy of Fallot (TOF), atrioventricular canal defect (AVSD), aortic stenosis (AS), truncus arteriosus, total anomalous pulmonary venous return (TAPVR)). Clusters 3 and 4 included severe CHD lesions (single ventricle, Ebstein’s anomaly, and hypoplastic left heart syndrome (HLHS)).

We binarized these clusters into normal (1) vs CHD (2-4) and calculated sensitivity, specificity, and accuracy of distinguishing normal vs CHD hearts. Sensitivity and specificity for CHD was 87% and 75%, respectively (Table 1).

**Table 1.**
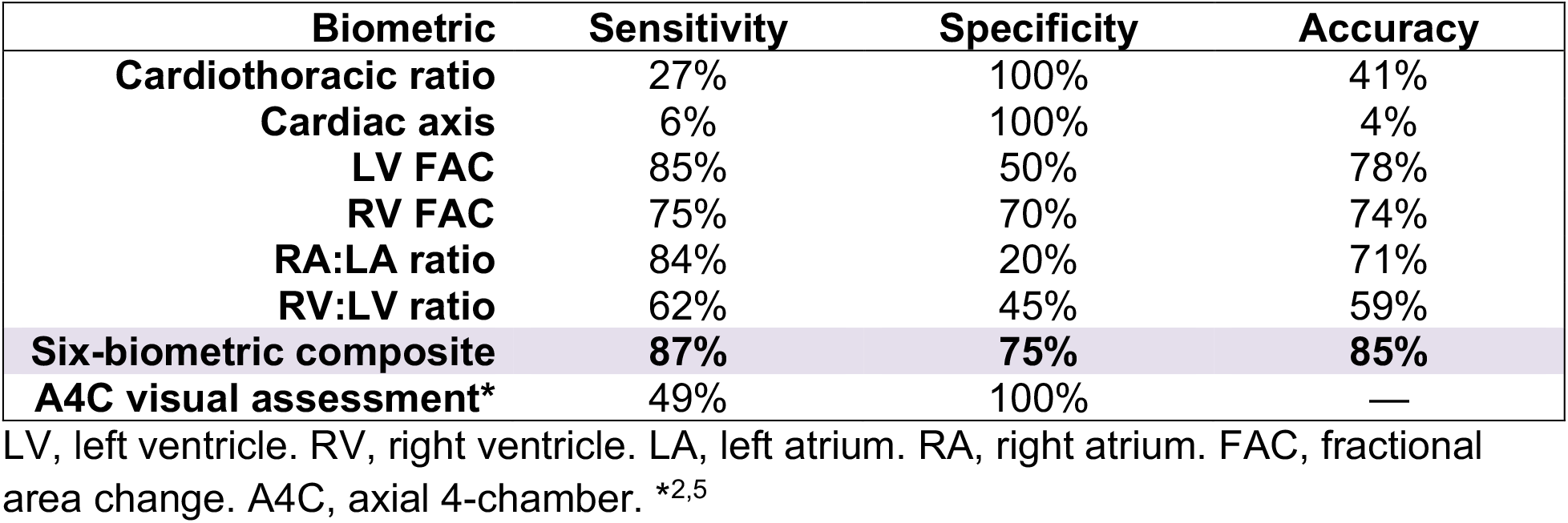
Performance of a composite biometric in distinguishing normal vs CHD hearts, compared to each individual biometric and to visual assessment as established by current literature.

Of the 11 false negatives, 8 belonged to lesions that often have a normal A4C (coarctation, TOF, dTGA). Other false negatives included two cases of single ventricle (SV) and one HLHS. For these SV cases, the ratio of the putative LV and RV segments were normal, even though there was no complete interventricular septum. Similarly, the false-negative HLHS was one where the LV was in fact dilated rather than hypoplastic, which again resulted in a normal RV:LV ratio.

Here, we demonstrate that a composite of six standard biometric has better sensitivity and accuracy for CHD than any one biometric alone and better than A4C visual assessment (Table 1)^2,5^. We focused on the biometrics described in order to test whether increased diagnostic accuracy is possible using measurements that are simple and already widely accepted, using the view most reliably obtained during screening. We used expert segmentation to remove the possibility that automatic segmentation (e.g., from a deep learning model^7^) could confound the findings.

Interestingly, there is improved accuracy even for lesions that traditionally have a normal A4C view. About 40% of CHD lesions are typically only appreciated in the outflow tract or 3-vessel views.^6^ 31 of our 105 fetal echocardiograms are lesions that typically have a normal A4C view. With our composite biometric, the sensitivity, specificity and accuracy for these lesions were 74%, 75% and 75% respectively.

Therefore, with composite analysis of all six biometrics, we can increase the utility of the A4C view even for these lesions. Given that A4C is the most reliably obtained cardiac view in the fetal screening ultrasound, the improvement in utility of the A4C view has profound screening implications. We do not expect for the composite biometric to replace overall clinical assessment, but rather to serve as an aid, similar to how single biometrics are currently used.

Despite its value, this study has several limitations. The size of the dataset analyzed is relatively small; however, it did include a comprehensive number of CHD lesions. Also, while there is improved diagnostic accuracy when using a composite of the six biometrics, this method is not perfect. The 11 false negative hearts had biometric features that calculated as normal with respect to the RV:LV ratio and CTR—which were important distinguishing features among clusters—despite being abnormal lesions. This shortcoming in fact raises the exciting possibilities that there may be additional, non-traditional biometrics that can be incorporated into the model for greater performance; these may come from the A4C view or, may incorporate other views such as the LVOT view. Testing additional non-standard biometrics within our model would be an interesting avenue for further study.

Now that we have shown the value-add of analyzing multiple biometrics within a composite model, advances in automation make multiple-biometric approaches increasingly practical. End-to-end automation (i.e., including automatic deep learning-based segmentation^7^) would eliminate manual tracing and allow interpretation of multiple biometrics at once in large data sets, thereby decreasing clinician burden and bringing enhanced biometric screening into practice. We look forward to further automating, refining, and scaling testing to prove clinical utility of composite biometrics in a larger population.

## Data Availability

Due to the vulnerable status of the patient population and waived consent, data will not be made available.

